# tDCS Improves Sleep in Adolescent Schizophrenia by Reorganizing Overlapping Brain Networks with Neurochemical and Transcriptomic Signatures: a Randomized Controlled Trial

**DOI:** 10.64898/2026.01.21.26344372

**Authors:** Yilin Huang, Bincan Xiong, Huashuang Zhang

## Abstract

**Background:** Chronic insomnia is a highly prevalent and disabling comorbidity in adolescent schizophrenia. Although schizophrenia has been increasingly conceptualized as a disorder of large-scale brain network disconnection, the mechanisms by which neuromodulation acts on overlapping system-level network architecture to improve sleep remain poorly understood. Here, we investigated the network, neurochemical, and transcriptomic mechanisms underlying transcranial direct current stimulation–based sleep improvement in adolescents with schizophrenia.

**Methods:** In a randomized, double-blind, sham-controlled trial, 78 adolescent schizophrenia with chronic insomnia received either active or sham transcranial direct current stimulation (tDCS). The anode was positioned over the left dorsolateral prefrontal cortex (DLPFC) and the cathode over the right DLPFC (20 sessions over 4 weeks). Insomnia Severity Index (ISI) and resting-state functional magnetic resonance imaging (fMRI) were assessed at baseline and post-treatment. We employed a multiscale analytical framework: Shannon-entropy diversity coefficients quantified overlapping system architecture; JuSpace toolbox assessed spatial correspondence with neurotransmitter maps; and the Allen Human Brain Atlas interrogated transcriptomic correlates.

**Results:** Active tDCS significantly improved insomnia symptoms compared to sham stimulation. This clinical improvement was accompanied by reorganization of overlapping network architecture involving 15 cortical regions with cross-network participation across attention, somatomotor, visual, and default-mode systems. Greater reduction in Shannon-entropy diversity coefficients was correlated with greater insomnia improvement. Spatially, such Shannon-entropy diversity coefficients changes aligned with the distribution of the dopaminergic transporter (DAT), metabotropic glutamate receptor 5 (mGluR5), and vesicular acetylcholine transporter (VAChT). At the molecular level, expression of 104 genes correlated with tDCS-sensitive regions, enriched for ionotropic glutamatergic signaling and AMPA/NMDA receptor complexes, and showed peak expression in cortical excitatory neurons during adolescence.

**Conclusions:** tDCS alleviates insomnia in adolescents with schizophrenia by restoring dysfunctional overlapping system architecture, a process spatially constrained by dopaminergic, glutamatergic, and cholinergic systems and supported by an adolescent-relevant glutamate gene network. Our findings reveal a multiscale pathway for neuromodulation and provide a biologically-grounded blueprint for developing sleep-focused personalized interventions in serious mental illness.

**Trial registration:** This study was registered in the Chinese Clinical Trial Registry (No. ChiCTR2600116100). Registered 5 January 2026 - Retrospectively registered, https://www.chictr.org.cn/showproj.html?proj=288899

## Background

Schizophrenia (SCZ) is a major psychiatric illness that affects around 1% of the global population [1]. Chronic insomnia is a highly prevalent comorbidity in SCZ and is associated with exacerbated cognitive deficits, emotional dysregulation, and impaired brain network function [2, 3]. Therefore, alleviating insomnia is not only a critical therapeutic goal in itself but may also represent a unique window into modulating the core pathophysiology of SCZ and improving overall functional outcomes.

Growing neuroimaging evidence conceptualizes SCZ as a disconnection syndrome, characterized by impaired interactions among large-scale brain networks [4–6]. However, most previous connectomic studies rely on non-overlapping network models, assuming each brain region belongs to a single network, thus neglecting the spatially overlapping organization of brain systems [7]. In contrast, emerging evidence suggests that many regions simultaneously contribute to multiple networks, forming an overlapping topology that supports cross-system integration [7]. These overlapping hubs typically show multimodal coactivation, hub-like structural features, and high dynamic flexibility. Their disruption has been linked to impaired large-scale integration in neuropsychiatric disorders [8, 9]. To characterize this architecture, we employed a recently developed information-theoretic measure—the Shannon entropy diversity coefficient—which quantifies the heterogeneity of a brain region’s cross-network connectivity profile [10, 11]. This framework provides an intuitive and quantitative way to capture system-level reorganization beyond traditional single-network models. Transcranial direct current stimulation (tDCS) is a promising non-invasive neuromodulation technique. Importantly, accumulating evidence suggests that tDCS can influence distributed brain networks rather than isolated regions, making it particularly suitable for probing system-level dysfunction in neuropsychiatric disorders. By inducing long-term potentiation (LTP)-like synaptic plasticity, tDCS can modulate cortical excitability and restore the excitation-inhibition (E/I) balance [12–14]. Its potential to normalize large-scale brain network communication makes it a compelling intervention for SCZ [15]. However, the precise mechanisms through which tDCS reshapes this overlapping network architecture—particularly to alleviate comorbid insomnia—remain unclear, limiting its targeted therapeutic optimization.

The organization and modulation of large-scale networks are constrained by the brain’s intrinsic biological architecture [16]. At the neurochemical level, functional heterogeneity is shaped by regional variations in neurotransmitter receptor densities [17]. A shared E/I imbalance, involving glutamatergic (e.g., mGluR5) and GABAergic systems, underpins both SCZ and insomnia [18–20] and influences local circuitry and global connectivity [21]. Our previous work suggests that neuromodulation can alleviate insomnia via GABAergic mechanisms [22]. To test if tDCS-induced network changes align with this neurochemical landscape, we used the JuSpace toolbox [23] which enables cross-scale analysis by integrating normative PET-derived neurotransmitter maps. This analysis bridges the gap between macroscopic network changes and the microscale neurotransmitter systems that directly govern synaptic efficacy and circuit excitability.

At the molecular level, spatial patterns of gene expression provide another foundational constraint. Transcriptome-connectome association studies link macroscale network organization to microscale gene expression gradients [24]. This approach has illuminated network dysfunction in disorders such as major depressive disorder and Parkinson’s disease[25, 26]. However, the relationship between gene expression and the overlapping network architecture in SCZ with comorbid insomnia—and how it is modulated by intervention—remains unexplored. Integrating these multi-scale perspectives (network, neurochemical, genomic) is critical for a unified mechanistic account of interventions like tDCS [27–29]. Yet no studies have examined how neurochemical constraints and transcriptomic gradients jointly shape tDCS-induced reorganization of overlapping system-level architecture in SCZ with insomnia.

To address these gaps, we conducted a randomized, double-blind, sham-controlled trial of tDCS in adolescents with SCZ and chronic insomnia. Within a multi-scale integrative framework, we tested three specific hypotheses: (i) tDCS induces reorganization of overlapping brain networks with regionally specific effects across functional systems, and this reorganization is associated with clinical improvement in insomnia symptoms. (ii) The baseline organization of overlapping network architecture would predict the degree of insomnia improvement following tDCS. (iii) The spatial pattern of tDCS-induced network reorganization in patients with SCZ is correlated with the distributions of disease-related neurotransmitters and expression maps of specific genes.

## Methods

### Study Design

We conducted a four-week, randomized, double-blind, controlled trial with two parallel arms: (A) active tDCS and (B) sham tDCS. All outcomes were assessed twice, at baseline and after completing a four-week intervention period. The Insomnia Severity Index (ISI) and resting-state functional magnetic resonance imaging (fMRI) were obtained at both time points. The study protocol was approved by the Ethics Committee of the Third People’s Hospital of Songzi. An overview of the analytic pipeline is presented in Fig. 1.

**Fig. 1.**
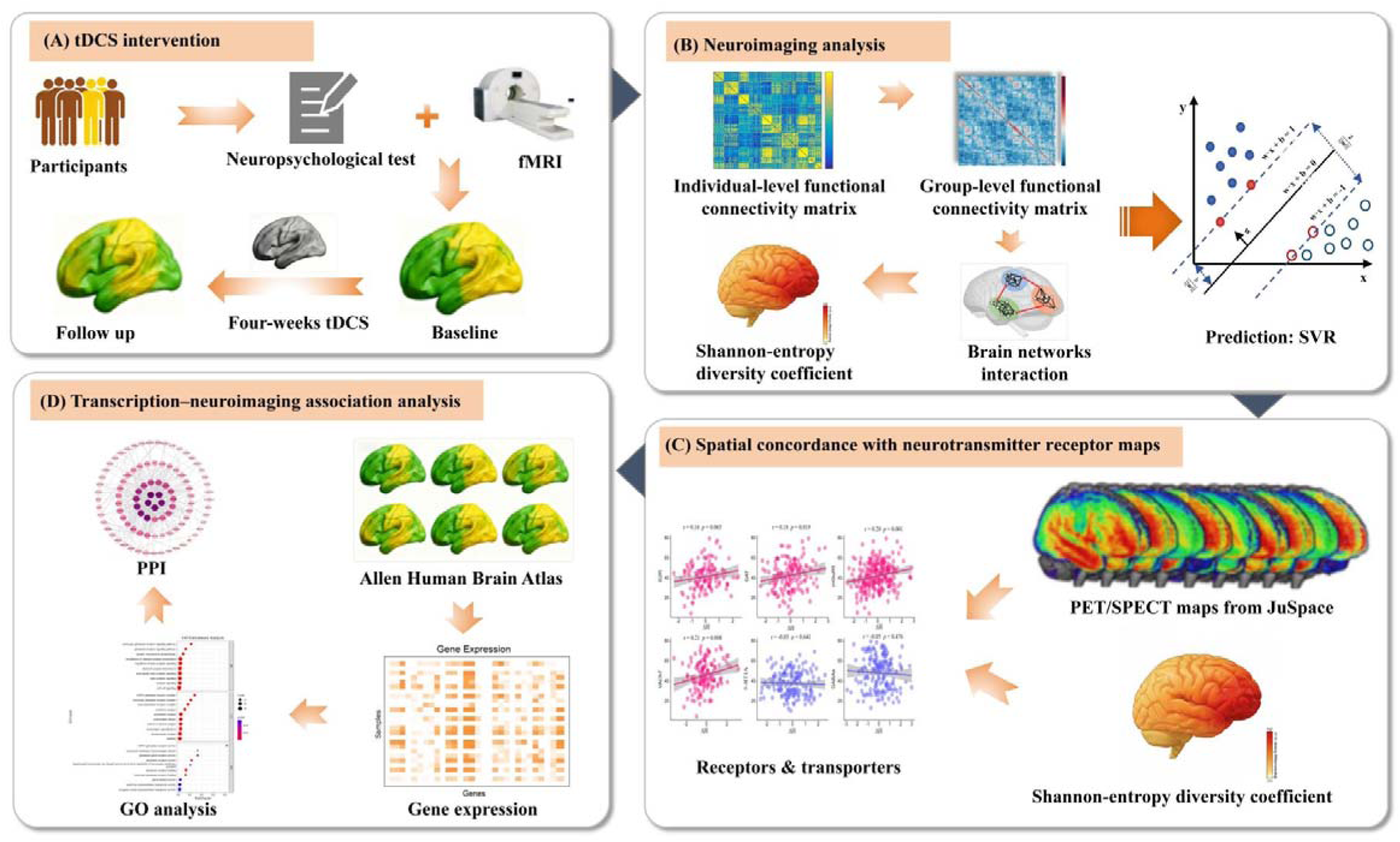
Study design and analysis pipeline. tDCS, transcranial direct current stimulation; fMRI: functional magnetic resonance imaging; 156 SVR: support vector regression; 157 GO: Gene Ontology; PPI: Protein protein interaction.

### Participants

Participants were recruited from the local community between March and July 2024 via advertisements, public talks and clinical referrals. Screening was performed by psychiatrists. Individuals meeting eligibility criteria were invited to an information session and provided written informed consent prior to enrolment. Inclusion criteria were: (1) age 12–18 years; (2) a DSM-5 diagnosis of schizophrenia; (3) DSM-5 diagnosis of chronic insomnia; (4) ISI score > 7; (5) illness duration (symptoms or diagnosis) ≥ 6 months before screening; (6) Positive and Negative Syndrome Scale (PANSS) total score ≥ 80 at both screening and baseline; and (7) if taking hypnotics, a stable dose for ≥ 4 weeks prior to baseline. Exclusion criteria included: (1) a primary DSM-5 diagnosis other than schizophrenia within three months prior to screening; (2) the presence of untreated sleep disorders, such as obstructive sleep apnoea, restless legs syndrome, or narcolepsy; (3) a past or current clinical presentation suggestive of delirium, dementia, amnestic syndromes, or other cognitive disorders; (4) psychotic symptoms attributable to an underlying medical condition or substance use; (5) hospitalization lasting 21 days or longer for an acute exacerbation of schizophrenia at baseline; (6) documented treatment resistance to at least two antipsychotic medications (with relapse related to non-adherence or substance use assessed at the investigator’s discretion); (7) a high risk of severe self-harm, suicide, or harm to others, including violent or homicidal behaviour; (8) a history of seizures, epilepsy, severe traumatic brain injury, or stroke; (9) unstable medical conditions involving hepatic, renal, cardiac, endocrine, neurological, or haematological systems that could increase study-related risk; (10) serious comorbid medical illnesses requiring continuous pharmacological treatment; (11) prior exposure to electroconvulsive therapy; (12) a positive drug screening result (with cannabis use evaluated on a case-by-case basis); and (13) current participation in another clinical trial.

### Sample size

Sample size was determined a priori using G*Power 3.1 [30] for a 2 × 2 repeated-measures ANOVA. Assuming a medium effect size (f = 0.25, ŋ² ≈ 0.06), α = 0.05, 90% power, two time points and a within-subject correlation of 0.5, the required total sample was 44 participants (22 per group). Anticipating a 20% dropout rate, we increased the target to 27 per group. To further protect power against higher-than-expected attrition, we ultimately enrolled 39 participants in each arm.

### Randomization and Blinding

Eligible participants were randomly allocated in a 1:1 ratio to active or sham tDCS. A blinded statistician generated the randomization sequence using computer-based random numbers. Participants and outcome assessors remained unaware of group allocation throughout the trial. Because of the practical requirements of stimulation delivery, the tDCS operator was not blinded. To maintain blinding, participants were informed that they would receive tDCS without specifying whether it was active or sham, and they were instructed not to discuss treatment details with blinded assessors.

### tDCS intervention

Participants received 20 stimulation sessions, delivered on weekdays over four consecutive weeks. Stimulation parameters followed Zhou et al. [31]. A tDCS-20A device delivered 2 mA of constant direct current. Current was applied through two 5×5 cm² saline-soaked sponge electrodes placed on rubber pads. Each session lasted 20 minutes. The anode was positioned over the left dorsolateral prefrontal cortex (DLPFC) (F3), and the cathode over the right DLPFC (F4) according to the International 10-20 EEG system [32, 33]. To minimize discomfort from abrupt current shifts, both onset and offset included 30-s ramp-up and ramp-down phases.

### Outcome measures

#### Primary outcome: fMRI

All MRI scans were acquired on a 3.0-T Philips scanner (Philips Medical Systems). Resting-state fMRI parameters were: repetition time (TR) = 2000 ms; echo time (TE) = 30 ms; slice thickness = 4.0 mm; flip angle = 90°; matrix = 64 × 64; 35 slices; voxel size = 3×3×3 mm³.

#### Secondary outcome: ISI

Insomnia was assessed using the ISI, a validated instrument designed to capture both nocturnal and daytime manifestations of insomnia [34]. Each item is rated on a 5-point scale ranging from 0 (“no difficulty”) to 4 (“severe difficulty”), yielding a total score between 0 and 28. Conventional thresholds classify scores of 0-7 as no clinically significant insomnia, 8-14 as subthreshold insomnia, 15-21 as moderate insomnia, and 22-28 as severe insomnia [35]. ISI is sensitive to changes following insomnia treatments and has demonstrated good reliability and validity [36].

### Neuroimaging data analysis

#### fMRI preprocessing

Resting-state fMRI data were preprocessed using the GRETNA toolbox [37] implemented in SPM12. To reduce initial magnetization instability, the first five volumes of each time series were discarded. Head motion was corrected via rigid-body realignment. Participants with excessive motion—defined as translation > 3 mm or mean framewise displacement (FD) > 0.5 mm—were excluded, resulting in the removal of two participants from the active tDCS group and one from the sham group. Following realignment, the time series underwent band-pass filtering (0.01–0.08 Hz) and nuisance regression in a unified linear model including 24 head-motion parameters, and mean signals from white matter, cerebrospinal fluid (CSF) and the whole brain[38]. White-matter and CSF masks were derived from SPM12 tissue probability maps (threshold = 0.9). Functional images were then normalized to the Montreal Neurological Institute (MNI) space using deformation fields from structural segmentation and resampled to 3-mm isotropic voxels.

#### Shannon-entropy diversity coefficient analysis

After preprocessing, we constructed individual functional connectivity (FC) matrices.

Mean time series were extracted from the 200 cortical parcels of the Schaefer-2018 atlas, which assigns each parcel to one of 17 functional systems. Pearson correlations were computed between all parcel pairs, and correlation coefficients were converted to z-scores using Fisher’s r-to-z transformation to improve normality and stabilize variance.

To obtain a robust modular structure for diversity estimation, the Louvain community detection algorithm (resolution parameter γ = 1) from the Brain Connectivity Toolbox was applied 1,000 times to each participant’s FC matrix, capturing variability in the stochastic optimization process. A co-assignment matrix was then constructed, summarizing how often each pair of nodes was assigned to the same community across iterations. Applying a consensus threshold of 0.5 to this matrix yielded a stable individual-level partition. Group-level consensus partitions were obtained by combining individual partitions within the active and sham tDCS groups separately.

Based on these group-level modules, we calculated the Shannon-entropy diversity coefficient (H) for each node. H, an information-theoretic index, quantifies how a node’s connectivity is distributed across modules, reflecting its participation in multiple overlapping systems. It is defined as:

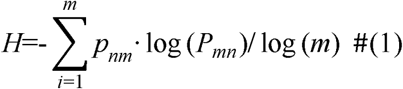

Where *P_nm_* represents the proportion of connection strength from node n to module m, and m is the total number of modules.

#### Identification of differential overlapping regions and links to clinical symptoms

To identify cortical regions whose system-level integration was altered by tDCS, we compared H values between the active and sham groups across pre- and post-treatment sessions. Regions exceeding a false discovery rate (FDR)–corrected threshold of p < 0.05 were defined as differential overlapping regions.

We then examined whether tDCS-related changes in diversity were associated with clinical improvement. For each significant region, treatment-specific change in H (ΔH) and changes in insomnia severity were computed as:

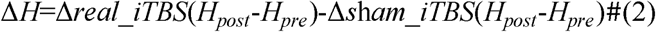

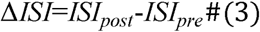

with more negative ΔISI values indicating greater improvement.

Partial correlations between ΔH and ΔISI were then computed across participants, controlling for age, sex, mean FD and baseline ISI. Statistical significance was assessed using 10,000-iteration permutation testing to obtain empirical p-values.

#### Baseline Overlap-Based Brain Signatures Predict tDCS Efficacy

Support vector regression (SVR) analyses were conducted using the LIBSVM toolbox. The diversity coefficients of the sleep related overlapping regions at baseline in the active tDCS group were used as features, and the change in ISI scores served as the target variable. Model performance was evaluated using leave-one-out cross-validation (LOOCV), and overall predictive accuracy was quantified by the root mean square error (RMSE) and pearson’s correlation coefficient.

#### Spatial concordance with neurotransmitter receptor maps

We evaluated the spatial alignment between tDCS-related functional reorganization and normative neurotransmitter distributions using JuSpace (v1.5) [23]. Six receptor or transporter systems were included: dopamine D2 receptor (D2R) [39], dopamine transporter (DAT) [40], metabotropic glutamate receptor 5 (mGluR5) [17], vesicular acetylcholine transporter (VAChT) [17], serotonin 1A receptor (5-HT1A) [41]and gamma-aminobutyric acid type-A receptors (GABA_A_) [38], all derived from PET templates. Multiple comparisons were controlled using false discovery rate (FDR) correction applied to the resulting p-values.

### Transcriptomic–neuroimaging association analysis

#### Preprocessing of gene expression

Transcriptomic data were from the Allen Human Brain Atlas (AHBA), which provides microarray-based expression profiles for over 20,000 genes across 3,702 tissue samples from six adult donors. Because right-hemisphere sampling is incomplete (only 4 of 6 donors), all transcriptomic analyses were restricted to the left hemisphere, following standard practice [42, 43]. This yielded a regional expression matrix for 100 left-hemisphere cortical parcels from the Schaefer-200 atlas. Preprocessing was performed using the abagen toolbox [42] and included: (1) probe filtering based on intensity > background in ≥ 50% of samples; (2) selection of the most differentially stable probe for each gene; (3) assignment of samples to the Schaefer-200 parcels using a 2-mm distance threshold; and (4) donor-wise z-score normalization to reduce inter-donor variability.

#### Association between gene expression and differential overlapping regions

To identify transcripts associated with tDCS-related changes in overlapping architecture, we examined the correspondence between regional gene expression and treatment-related changes in H within regions showing significant overlap alterations between active and sham conditions. For each of these regions, we extracted AHBA expression values and computed cross-regional Pearson correlations between gene expression and ΔH.

To account for spatial autocorrelation and multiple testing, a two-stage procedure was used. First, for each gene, a spatial permutation test (10,000 spin rotations of the brain map using the spin_test function in the neuromaps toolbox) was applied to derive spatially corrected p-values. Rotations preserved hemisphere assignments, and parcels overlapping the medial wall were excluded to maintain realistic distance relationships. Second, FDR correction (p < 0.05) was applied to the spatially corrected p-values. Only genes surviving both steps were retained as significantly associated with tDCS-induced overlap changes.

#### Disease-related differential expression analysis

To assess disorder specificity of the imaging-derived genes, we extracted logL fold-change (logLFC) values from the cross-disorder transcriptomic dataset of Gandal et al. [44], which includes autism spectrum disorder (ASD), major depressive disorder (MDD), SCZ, bipolar disorder (BD), alcoholism and inflammatory bowel disease (IBD). For each disorder, we calculated spearman correlations between the imaging–gene association coefficients and the corresponding logLFC values. Statistical significance was evaluated using 10,000-iteration permutation tests with FDR correction (p < 0.05). A significant or oppositely directed correlation specific to a given disorder was interpreted as evidence of disease-specific transcriptional coupling.

#### Gene enrichment analysis

Genes identified in the association analyses were subjected to functional enrichment. We used the ToppGene platform (https://toppgene.cchmc.org/) [45] for annotation and prioritization. Gene Ontology (GO) enrichment was conducted across biological process, cellular component and molecular function categories. Tissue, cell-type and developmental stage enrichment were then examined using Tissue-Specific Expression Analysis (TSEA; http://genetics.wustl.edu/jdlab/tsea/) and the Cell Type-Specific Expression Analysis (CSEA) tool (http://genetics.wustl.edu/jdlab/csea-tool-2/). Specificity index probabilities (P^SI^ = 0.05, 0.01 and 0.001) quantified the likelihood of preferential expression in particular tissues, cell types or developmental periods [46].

#### Protein–protein interaction (PPI) analysis

To explore interactions among the associated genes, we constructed a protein–protein interaction network using STRING v12.0. Nodes within the top 30% of degree centrality and a minimum confidence score of 0.45 were designated as hub genes. The developmental expression patterns of these hub genes were further explored using the Human Brain Transcriptome Database.

## Results

### Participant characteristics

Baseline demographic characteristics of participants in the active and sham tDCS groups are summarized in Table 1, with no significant differences observed between groups.

**Table 1.**
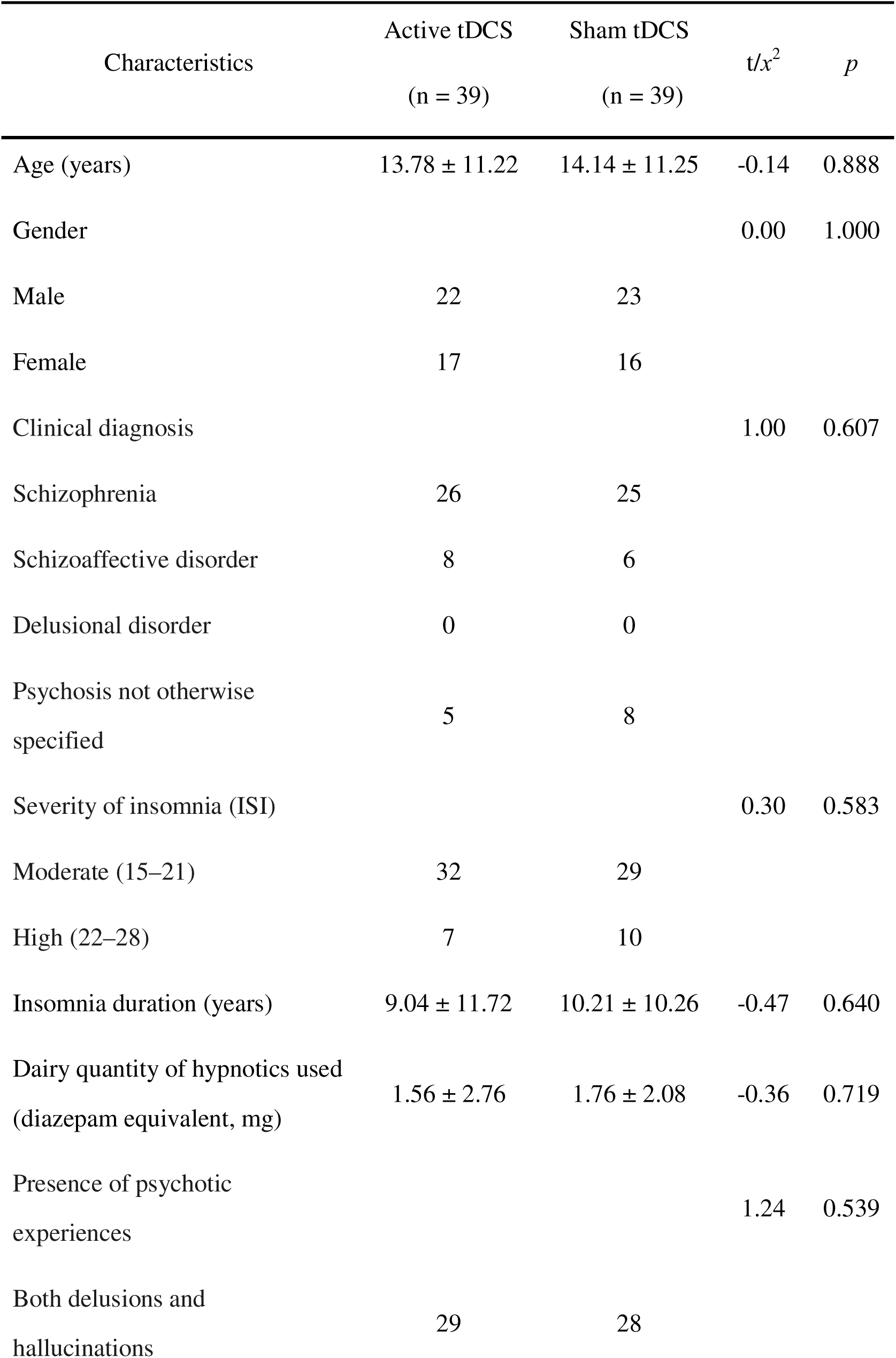
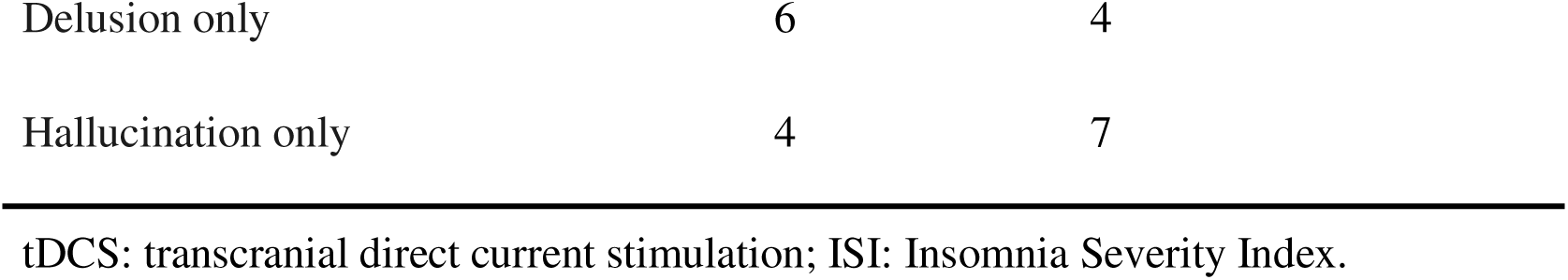
Demographics and clinical characteristics.

| Characteristics | Active tDCS<br>(n = 39) | Sham tDCS<br>(n = 39) | $t/x^2$ | $p$ |
| --- | --- | --- | --- | --- |
| Age (years) | 13.78 ± 11.22 | 14.14 ± 11.25 | -0.14 | 0.888 |
| Gender |  |  | 0.00 | 1.000 |
| Male | 22 | 23 |  |  |
| Female | 17 | 16 |  |  |
| Clinical diagnosis |  |  | 1.00 | 0.607 |
| Schizophrenia | 26 | 25 |  |  |
| Schizoaffective disorder | 8 | 6 |  |  |
| Delusional disorder | 0 | 0 |  |  |
| Psychosis not otherwise specified | 5 | 8 |  |  |
| Severity of insomnia (ISI) |  |  | 0.30 | 0.583 |
| Moderate (15–21) | 32 | 29 |  |  |
| High (22–28) | 7 | 10 |  |  |
| Insomnia duration (years) | 9.04 ± 11.72 | 10.21 ± 10.26 | -0.47 | 0.640 |
| Dairy quantity of hypnotics used<br>(diazepam equivalent, mg) | 1.56 ± 2.76 | 1.76 ± 2.08 | -0.36 | 0.719 |
| Presence of psychotic experiences |  |  | 1.24 | 0.539 |
| Both delusions and hallucinations | 29 | 28 |  |  |

|  |  |  |
| --- | --- | --- |
| Delusion only | 6 | 4 |
| Hallucination only | 4 | 7 |

tDCS: transcranial direct current stimulation; ISI: Insomnia Severity Index.

### Clinical outcome

Changes in ISI in both groups of active tDCS and sham tDCS are presented in Additional file 1: Fig. S1. Paired samples t-tests revealed that the scores of ISI in the active tDCS group were remarkably reduced by 8.76 points (t = 4.05, p < 0.001, Cohen’s d = 0.90), but no remarkable changes existed in the sham tDCS group. A univariate analysis of covariance (ANCOVA) was conducted to compare post-treatment ISI scores between groups, demonstrating significantly greater improvement in insomnia symptoms in the active tDCS group relative to controls (F = 14.10, p < 0.001).

### Network organization differences before and after tDCS

Before computing the Shannon-entropy diversity coefficient, we delineated 17 large-scale cortical networks, comprising the control (Cont A–C), default mode (Default A–C), dorsal attention (DorsAttn A–B), limbic (Limbic A–B), salience/ventral attention (SalVentAttn A–B), somatomotor (SomMot A–B), temporal–parietal (TempPar), visual central (VisCent), and visual peripheral (VisPeri) systems. To facilitate visualization, each network within the Schaefer-200 cortical parcellation was color-coded according to its spatial correspondence with the canonical 17-network atlas. Compared with the sham group, participants in the active tDCS group showed pronounced changes in functional connectivity across multiple cortical nodes. In contrast, the sham tDCS group showed no appreciable differences in network configuration between the pre- and post-treatment sessions.

### Differences in overlapping brain architecture between groups

To map alterations in overlapping system organization across the 17 large-scale cortical networks, Shannon-entropy diversity coefficients were computed for every cortical region at both pre- and post-treatment assessments in the active and sham tDCS groups. Group comparisons revealed 15 cortical regions that showed significant treatment-related changes in the active tDCS group relative to the sham group (*p* < 0.05, FDR corrected). These regions were predominantly distributed within the dorsal attention/salience–ventral attention network (DorsAttn/SalVentAttn; n = 5), the somatomotor network (SomMot; n = 6), the visual network (n = 1) and default network (n = 3) (Fig. 2).

**Fig. 2.**
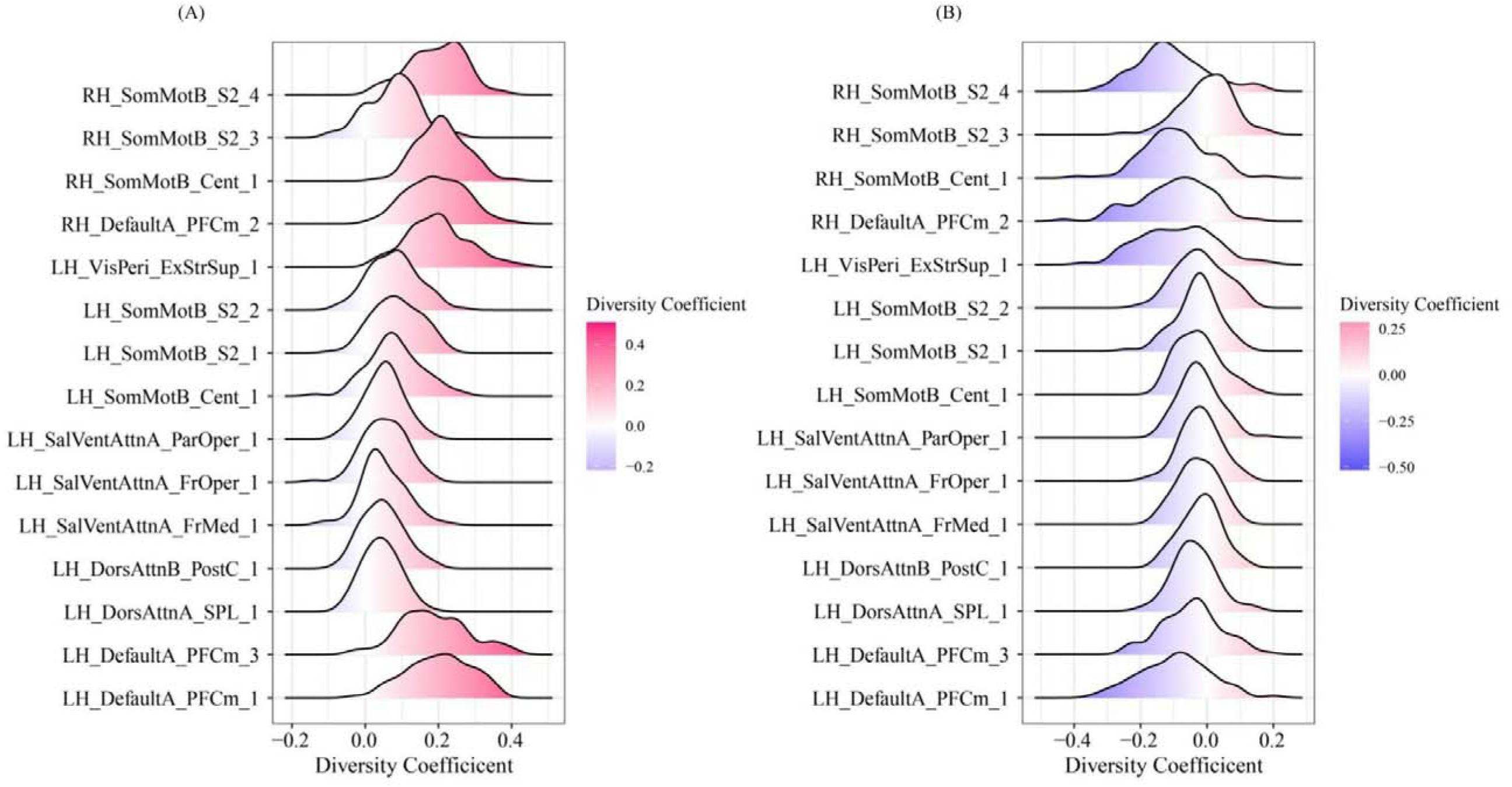
Changes in the Shannon-entropy diversity coefficient in 15 differential brain overlapping regions after four weeks of tDCS intervention. Differences in the sity coefficients before and after treatment were observed in the (**A**) active tDCS group and (**B**) sham tDCS group.

### Prediction of tDCS efficacy using baseline Shannon-entropy diversity coefficients

Analysis of behavioral relevance identified 7 out of 15 brain regions whose baseline Shannon-entropy diversity coefficients were significantly associated with insomnia severity (pL<L0.05). These regions predominantly belonged to the somatomotor and visual networks. We used the baseline entropy values of these 7 regions as features in a SVR model to predict the change in ISI scores after 4 weeks of treatment. These regions were primarily distributed within the somatomotor and visual networks. baseline Shannon-entropy diversity coefficients from the 7 regions were then entered as predictors in a SVR model to estimate changes in ISI scores after four weeks of treatment. The model demonstrated significant predictive accuracy, with a strong correlation between predicted and observed improvement in sleep symptoms (rL=L0.62, pL=L0.002) (Additional file 1: Fig. S2).

### Correlation between Shannon-entropy diversity coefficients and neurotransmitter profiles

The Shannon-entropy diversity coefficients showed significant correlations with 3 neurotransmitter receptor profiles: DAT (r = 0.18, *p* = 0.019, FDR corrected), mGluR5 (r = 0.20, *p* = 0.001, FDR corrected) and VAChT (r = 0.21, *p* = 0.008, FDR corrected). No significant associations were observed for the remaining neurotransmitter systems, including D2R (r = 0.16, *p* = 0.065), 5-HT1A (r = −0.03, *p* = 0.641) and GABA_A_ (r = −0.05, *p* = 0.476) (Fig. 3A-F).

**Fig. 3.**
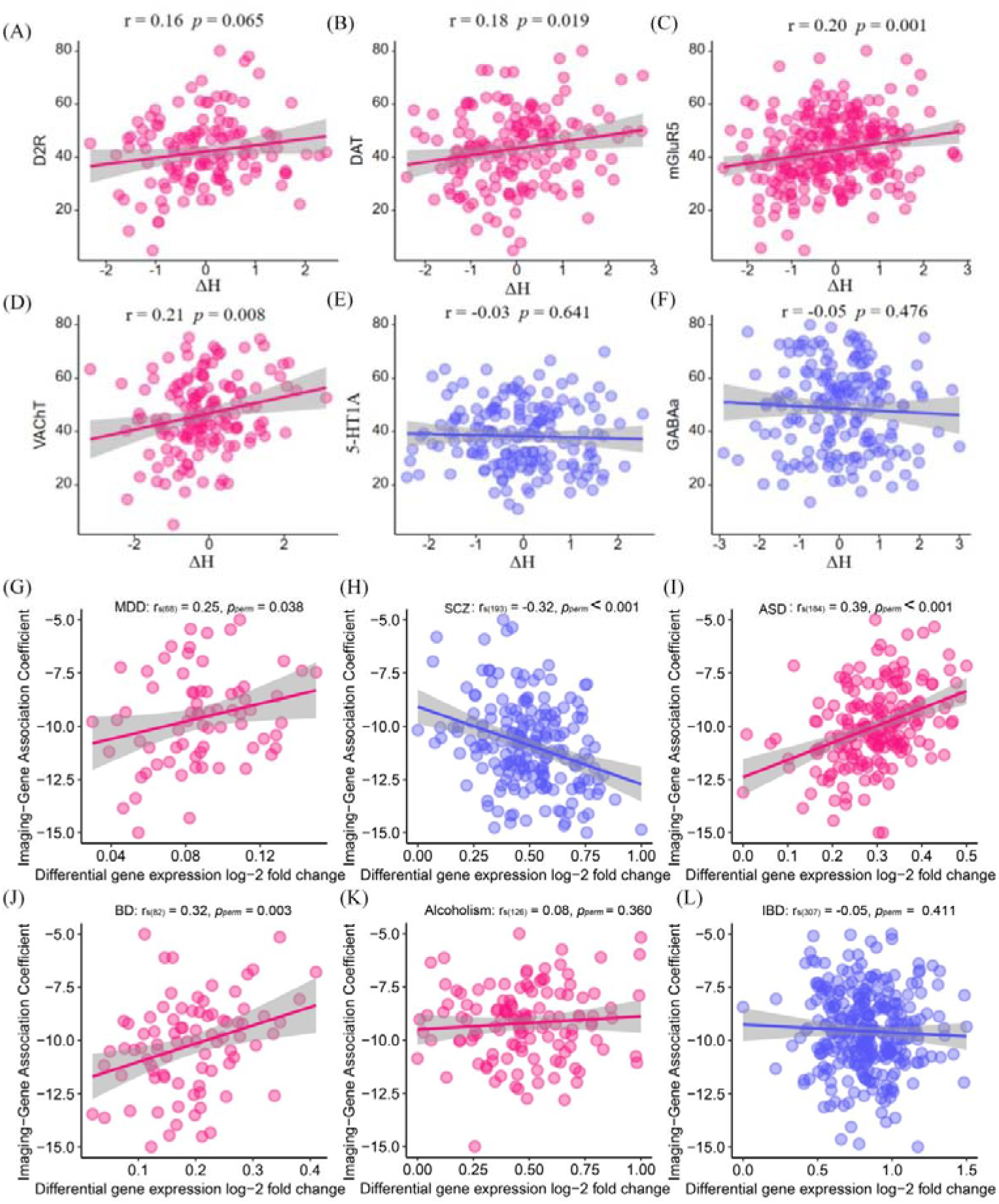
Multiscale associations between network alterations, neurotransmitter distribution, and disorder-related gene expression. (**A-F**) Spatial correlations between changes in the Shannon-entropy diversity coefficient (ΔH) and normative neurotransmitter receptor/transporter density maps, including D2R, DAT, mGluR5, VAChT, 5-HT1A, and GABA_A_. (**G-L**) Associations between imaging-gene association coefficients and disorder-related differential gene expression (log_2_ fold change) derived from cross-disorder transcriptomic signatures reported by Gandal et al. [44]. D2R, dopamine D2 receptor; DAT, dopamine transporter; mGluR5, metabotropic glutamate receptor 5; VAChT, vesicular acetylcholine transporter; 5-HT1A, serotonin 1A receptor; GABA_A,_ gamma-aminobutyric acid type A receptors; MDD, major depressive disorder; ASD, autism spectrum disorder; SCZ, schizophrenia; BD, bipolar disorder; IBD, inflammatory bowel disease.

### Association between differential overlapping brain architecture and gene expression

Normalized transcriptional data for over 16,000 genes were obtained from 3,121 tissue samples in the AHBA following reannotation and refined probe selection [42]. Gene expression values were mapped to cortical parcels to generate a left-hemisphere regional expression matrix. Spatial correlations were then computed between regional gene expression profiles and changes in the Shannon-entropy diversity coefficient. In the active tDCS group, 104 genes were identified (p < 0.05, FDR corrected; Additional file 1:Table S1) whose expression patterns significantly aligned with the spatial distribution of tDCS-induced alterations in the overlapping brain system.

### Disease-related differential expression analysis

We identified 195 genes that were shared between our gene set and those reported by Gandal et al. [44]as significantly dysregulated in postmortem SCZ case-control brain tissue. Moreover, the spatial association coefficients of these genes showed a significant negative correlation with their SCZ-related expression alterations (r_s(193)_ = −0.32, adjusted *p*_perm_ < 0.001, FDR corrected). This association was specific to SCZ, as it was not observed in the other five disorders: MDD (r_s(68)_ =0.25, adjusted *p*_perm_ = 0.038, FDR corrected), ASD (r_s(184)_ = 0.39, adjusted *p*_perm_ < 0.001, FDR corrected), BD (r_s(82)_ = 0.32, adjusted *p*_perm_ = 0.003, FDR corrected), alcoholism (r_s(126)_ = 0.08, adjusted *p*_perm_ = 0.360, FDR corrected), and IBD (r_s(307)_ = −0.05, adjusted *p*_perm_ = 0.411, FDR corrected) (Fig. 3G-L).

### Gene functional enrichment

To elucidate the biological relevance of the tDCS-sensitive gene set, we performed GO enrichment analysis. GO enrichment results revealed significant clustering within glutamatergic synaptic signaling pathways. A global overview of ontology categories demonstrated that these pathways accounted for the major proportion of enriched terms, while quantitative analysis further identified the ionotropic glutamate receptor signaling pathway, AMPA receptor complex, and glutamate receptor activity as the most enriched GO terms (Fig. 4A).

**Fig. 4.**
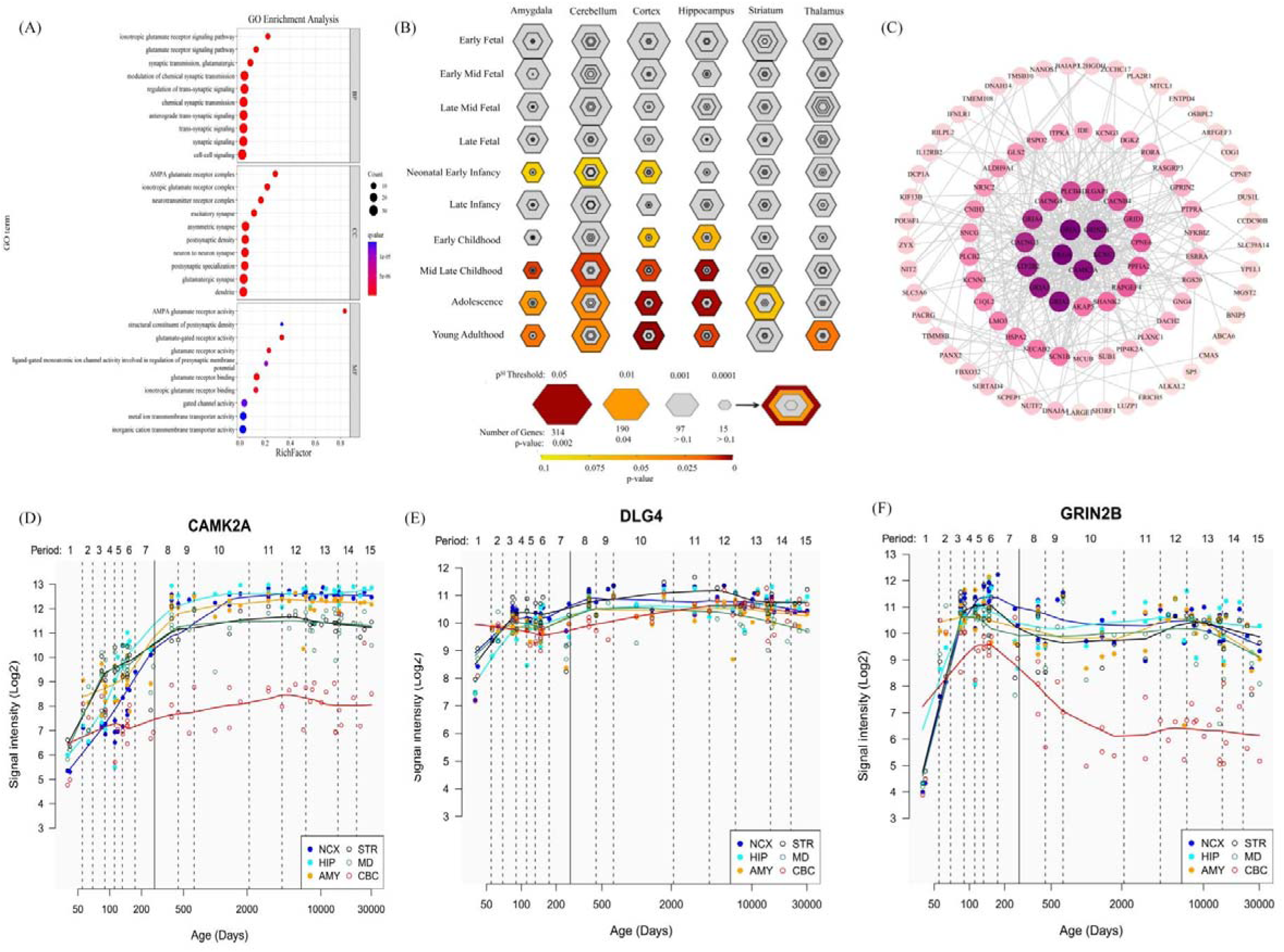
Transcriptomic features associated with alterations in the Shannon-entropy diversity coefficient. (**A**) GO enrichment analysis of genes associated with changes in the Shannon-entropy diversity coefficient, highlighting enrichment in glutamatergic synaptic signaling pathways. (**B**) Spatiotemporal enrichment of these genes across brain regions and developmental stages using the p^SI^ framework, showing preferential enrichment in cortical and hippocampal regions from late childhood through adolescence. (**C**) PPI network of the identified genes, with CAMK2A, DLG4, and GRIN2B emerging as central hub genes. (**D-F**) Developmental expression trajectories of (**D**) CAMK2A, (**E**) DLG4, and (**F**) GRIN2B across multiple brain regions, illustrating developmentally regulated expression patterns. GO, Gene Ontology; PPI, Protein–protein interaction.

### Tissue, cell type-and temporal-specific expression

Tissue specificity analyses identified the brain as the primary site of expression for these genes (Additional file 1: Fig. S3A). At the cellular level, significant enrichment was observed in cortical excitatory neuronal populations as well as striatal D1+ spiny projection neurons (Additional file 1: Fig. S3B). Temporal expression analysis revealed that the cortex exhibited the most prominent enrichment, spanning multiple critical developmental windows from early childhood through mid-to-late childhood, adolescence and young adulthood. Additional enrichment was also observed in the cerebellum and hippocampus, though with a narrower developmental distribution, while weaker signals appeared in the amygdala and striatum (Fig. 4B)

### PPI networks and hub genes

A PPI network (Fig. 4C) constructed from the identified genes revealed a highly interconnected core centered on CAMK2A, DLG4, GRIN2B, and their interacting partners. The spatiotemporal expression trajectories of these hub genes further demonstrated pronounced developmental regulation, characterized by increasing postnatal expression and sustained high levels across cortical and hippocampal regions from adolescence into young adulthood (Fig. 4D-F).

## Discussion

Chronic insomnia is a highly disabling and undertreated comorbidity in adolescents with SCZ, which exacerbates symptom severity, cognitive impairment, and adverse long term outcomes. Therefore, elucidating the neurobiological mechanisms underlying sleep improvement in this population is crucial for the development of targeted interventions. In this randomized controlled trial, we demonstrate that a four-week course of tDCS applied to the DLPFC not only significantly alleviates insomnia symptoms, but also drives reorganization of overlapping large-scale brain systems, with the most pronounced changes occurring in the dorsal attention/salience–ventral attention, somatomotor, visual, and default mode networks. JuSpace analysis further revealed that the effects of tDCS are spatially constrained by the distributions of DAT, mGluR5, and VAChT systems—three neurotransmitter systems deeply implicated in the neurobiology of adolescent SCZ with chronic insomnia. Transcriptomic profiling additionally uncovered convergent gene-expression patterns across tDCS-sensitive regions. These findings provide a multiscale integrative perspective—from systems-level networks to neurochemical architecture and molecular expression—for understanding how neuromodulation improves sleep in adolescents with SCZ.

The changes induced by tDCS were most prominent in cortical regions belonging to the dorsal attention/salience-ventral attention networks, the somatomotor network, the visual network, and the default mode network. Notably, the degree of normalization in this overlapping network architecture, quantified using the Shannon-entropy diversity coefficient, was positively associated with the extent of improvement in insomnia symptoms. This suggests that enhanced cross-system integration among key large-scale networks may support more restorative sleep-related functions. This finding is consistent with the notion that overlapping hubs mediate flexible inter-system information routing [47], and challenges traditional non-overlapping network models that conceptualize the brain as a set of discrete, unifunctional modules [48,49]. More specifically, abnormal interactions between the default-mode network and the dorsal attention network have been implicated in sleep-onset difficulties in insomnia. It is thus plausible that the tDCS-induced rebalancing of engagement across these overlapping networks contributes to fostering a neural state more conducive to sleep initiation.

Our JuSpace analysis further revealed that tDCS-induced network modulation showed high spatial concordance with the distributions of dopaminergic (DAT), glutamatergic (mGluR5), and cholinergic (VAChT) systems. This strongly suggests that the effects of tDCS are not diffuse or nonspecific, but are instead constrained by the brain’s intrinsic chemoarchitectural organization. In support of this, recent evidence demonstrates that glutamatergic—but not cholinergic—neurons in the basal forebrain can trigger activity-dependent adenosine release, a critical molecular mediator of sleep pressure and homeostatic regulation of the sleep–wake cycle [50]. This finding links excitatory neuronal activity during wakefulness to the build-up of sleep-inducing signals, thereby providing a mechanistic bridge between glutamatergic tone, sleep homeostasis, and cortical excitability. In this context, tDCS-induced modulation of glutamatergic pathways may enhance endogenous sleep pressure signaling and facilitate sleep recovery in individuals with chronic insomnia. Thus, our results support the hypothesis that tDCS exerts its effects by acting on specific network topologies defined by these receptor gradients [17], thereby providing a chemoarchitectural basis for developing more precisely targeted neuromodulation strategies.

Our transcriptomic analysis revealed that tDCS modulates a developmentally sensitive molecular program centered on glutamatergic synapses, with prominent enrichment of AMPA receptor subunits (GRIA1–4) and NMDA receptor subunits (GRIN2B). These molecules are key mediators of activity-dependent synaptic strengthening and receptor compositional plasticity, respectively [51, 52]. This program exhibits clear spatiotemporal specificity, as the target genes show high expression in cortical excitatory neurons and striatal D1+ spiny projection neurons, with expression peaking during adolescence. This developmental period is characterized by both heightened synaptic remodeling and increased vulnerability to psychiatric disorders[53]. Such a pattern suggests that tDCS administered during adolescence may more effectively engage fronto-striatal circuits that are in a state of elevated plastic potential.

At the molecular level, the membrane insertion of GluR1-containing AMPA receptors is considered a fundamental mechanism of LTP [54]. Meanwhile, the dynamic regulation of GRIN2B-containing NMDA receptors is critical for cortical plasticity and has been identified as a central pathological mechanism in schizophrenia [55,56]. This view is supported by preclinical evidence indicating that reduced NMDAR function can induce schizophrenia-like behavioral phenotypes, which are reversible with pharmacological interventions targeting dopaminergic and serotonergic systems [57,58]. Therefore, tDCS-induced changes in gene expression may contribute to compensating for underlying NMDAR hypofunction in at-risk individuals.

The enhancement of NMDAR function promotes calcium influx, which activates downstream pathways such as the CREB–BDNF signaling cascade. This process facilitates synaptic remodeling and increases the excitability of specific neuronal ensembles [59, 60]. We suggest that these molecular and cellular effects provide a mechanistic foundation for the large-scale network reorganization observed in our neuroimaging data, particularly within circuits such as the dorsal attention and salience networks. Improved network integration may ultimately stabilize the neural dynamics that regulate sleep–wake transitions. For instance, this could occur through reduced activity in the default mode network, which has been associated with pathological rumination.

The most important translational implication of this study is that it establishes chronic insomnia as a mechanistically defined and modifiable treatment target in adolescents with SCZ. Improving sleep not only alleviates a core comorbidity, but may, through the aforementioned multiscale mechanisms—namely, neurochemically constrained network reorganization and developmentally specific synaptic plasticity—serve as a “clinical entry point” for promoting broader recovery in cognition and everyday functioning. Our findings provide a conceptual framework for future precision neuromodulation, in which individualized selection of tDCS targets and parameters may be guided by each patient’s neurotransmitter receptor distribution profile or associated gene-expression signatures. This directly echoes the aims of the present Collection, which calls for mechanistic understanding of sleep–mental health comorbidity and the development of mechanism-based, youth-focused interventions.

Several limitations of this study should be acknowledged. First, AHBA is based on neurotypical adult donors and has limited sampling of the right hemisphere, which may introduce bias when integrating these transcriptomic data with disease-specific neuroimaging findings. Second, although interest in overlapping system-level architecture is growing, analytic methods for characterizing this organization—such as the Shannon-entropy diversity coefficient used in the present study—are still in an early stage of development and require validation in larger and independent samples. Finally, although our multimodal findings converge on a biologically plausible pathway, the direct causal chain linking network-level and molecular changes to sleep improvement has yet to be established. Future research could combine tDCS with PET to directly verify its modulatory effects on specific receptor systems, validate the predictive utility of network-based biomarkers in prospective large-scale cohorts, and explore interventions that temporally align tDCS with specific sleep–wake or circadian phases, in order to further test and refine the mechanistic model proposed here.

## Conclusions

In summary, this study provides multiscale evidence that tDCS improves sleep in adolescents with SCZ through a specific pathway constrained by neurochemical architecture and involving adolescence-relevant glutamatergic synaptic plasticity gene programs. These findings not only clarify the mechanisms by which tDCS exerts its effects, but also lay an empirical foundation for developing neuroscience-informed precision sleep interventions for adolescents with SCZ. This approach may pave the way for a new generation of neurobiologically-informed therapies that move beyond symptom management to target core pathophysiology.

## Abbreviations

tDCS: transcranial direct current stimulation
DLPFC: dorsolateral prefrontal cortex
ISI: Insomnia Severity Index
fMRI: functional magnetic resonance imaging
SCZ: Schizophrenia
LTP: long-term potentiation
E/I: excitation-inhibition
PANSS: Positive and Negative Syndrome Scale
TR: repetition time
TE: echo time
FD: framewise displacement
CSF: cerebrospinal fluid
MNI: Montreal Neurological Institute
FC: functional connectivity
FDR: false discovery rate
SVR: support vector regression
LOOCV: leave-one-out cross-validation
RMSE: root mean square error
D2R dopamine: D2 receptor
DAT: dopamine transporter
mGluR5: metabotropic glutamate receptor 5
VAChT: vesicular acetylcholine transporter
5-HT1A: serotonin 1A receptor
GABA_A_: gamma-aminobutyric acid type-A receptors
FDR: false discovery rate
AHBA: Allen Human Brain Atlas
log-JFC: log-J fold-change
ASD: autism spectrum disorder
MDD: major depressive disorder
BD: bipolar disorder
IBD: inflammatory bowel disease
GO: Gene Ontology
TSEA: Tissue-Specific Expression Analysis
CSEA: Cell Type-Specific Expression Analysis
PPI: Protein–protein interaction

## Supplementary Information

Additional file 1 (Additional file 1.pdf): Tables S1 and Fig. S1-S3. Table S1 List of the genes identified in this study. Fig. S1 ISI scores at pre-tDCS and post-tDCS of the two groups. Fig. S2 SVR analysis revealed a significant correlation between these baseline diversity coefficients and actual sleep improvement, indicating that the baseline brain overlapping system can predict tDCS treatment effects on sleep. Fig. S3 Tissue- and cell-type enrichment analyses reveal that SCZ-related genes are predominantly enriched in brain tissue, with pronounced signals in cortical neurons, indicating a neuron- and cortex-centered molecular architecture underlying SCZ-related network alterations.

## Declarations

### Ethics approval and consent to participate

Ethics approval was granted by Third People’s Hospital of Songzi (202404XA22). This study was conducted in accordance with the ethical principles of the Declaration of Helsinki. All participants provided written informed consent.

### Consent for publication

Not applicable.

### Data availability

PET-derived neurotransmitter distribution maps are accessible via the JuSpace Toolbox (https://github.com/juryxy/JuSpace) within MATLAB R2024b. Postmortem gene expression profiles are available from the Allen Human Brain Atlas (https://help.brain-map.org). Gene expression data from the Allen Human Brain Atlas were preprocessed using the abagen toolbox (https://github.com/rmarkello/abagen) within Python 3.12.7. Gene enrichment analysis was performed using Metascape (https://metascape.org/gp/index.html). Brain mapping images were visualized with MRIcron (https://www.nitrc.org/projects/mricron). The datasets generated and/or analyzed during the current study are available from the corresponding author upon reasonable request. Due to the sensitive nature of the clinical data and restrictions of informed consent, participant-level data are not publicly available.

### Authors’ contributions

YH: Writing-original draft, Conceptualization, Investigation, Formal analysis, Data curation, Funding acquisition, Resources. BX: Data curation, Visualization, Software, Methodology, Investigation, Supported data analyses, Resources. HZ: Editing & Validation. All authors read and approved the final manuscript.

## Data Availability

https://help.brain-map.org

https://github.com/rmarkello/abagen

https://metascape.org/gp/index.html

https://www.nitrc.org/projects/mricron

## Acknowledgement

We would like to thank all participants who took part in our study.

## Author details

^1^Department of Medicine, Foshan University, Foshan 528000, China.

## Statements (Author contributions & Ethic statement)

### Contributors

Y.H. and B.X. conceived and designed the study; H.Y., B.X. and H.Z. collected the data and performed the analysis; Y.H. and B.X. conducted the investigation and provided resources; Y.H. wrote the original draft; H.Z. reviewed, edited and validated the manuscript. All authors read and approved the final version.

## References

1. McCutcheon RA, Reis Marques T, Howes OD. Schizophrenia-an overview. JAMA Psychiatry. 2020;77:201–10.

2. Manoach DS, Stickgold R. Abnormal sleep spindles, memory consolidation, and schizophrenia. Annu Rev Clin Psychol. 2019;15:451–79.

3. Reeve S, Sheaves B, Freeman D. The role of sleep dysfunction in the occurrence of delusions and hallucinations: A systematic review. Clin Psychol Rev. 2015;42:96–115.

4. Fornito A, Zalesky A, Breakspear M. The connectomics of brain disorders. Nat Rev Neurosci. 2015;16:159–72.

5. Friston KJ. The disconnection hypothesis. Schizophr Res. 1998;30:115–25.

6. Pettersson-Yeo W, Allen P, Benetti S, McGuire P, Mechelli A. Dysconnectivity in schizophrenia: Where are we now? Neurosci Biobehav Rev. 2011;35:1110–24.

7. Faskowitz J, Esfahlani FZ, Jo Y, Sporns O, Betzel RF. Edge-centric functional network representations of human cerebral cortex reveal overlapping system-level architecture. Nat Neurosci. 2020;23:1644–54.

8. Jo Y, Zamani Esfahlani F, Faskowitz J, Chumin EJ, Sporns O, Betzel RF. The diversity and multiplexity of edge communities within and between brain systems. Cell Rep. 2021;37:110032.

9. Gu Y, Li L, Zhang Y, Ma J, Yang C, Xiao Y, et al. The overlapping modular organization of human brain functional networks across the adult lifespan. Neuroimage. 2022;253:119125.

10. Martínez García SJ, Padilla Longoria P. Analysis of shannon’s entropy to contrast between the embodied and neurocentrist hypothesis of conscious experience. Biosystems. 2024;246:105323.

11. Rubinov M, Sporns O. Weight-conserving characterization of complex functional brain networks. Neuroimage. 2011;56:2068–79.

12. Barron HC, Vogels TP, Emir UE, Makin TR, O’Shea J, Clare S, et al. Unmasking latent inhibitory connections in human cortex to reveal dormant cortical memories. Neuron. 2016;90:191–203.

13. Bove F, Angeloni B, Sanginario P, Rossini PM, Calabresi P, Di Iorio R. Neuroplasticity in levodopa-induced dyskinesias: An overview on pathophysiology and therapeutic targets. Prog Neurobiol. 2024;232:102548.

14. Nitsche MA, Kuo MF, Karrasch R, Wächter B, Liebetanz D, Paulus W. Serotonin affects transcranial direct current-induced neuroplasticity in humans. Biol Psychiatry. 2009;66:503–8.

15. Mondino M, Jardri R, Suaud-Chagny MF, Saoud M, Poulet E, Brunelin J. Effects of fronto-temporal transcranial direct current stimulation on auditory verbal hallucinations and resting-state functional connectivity of the left temporo-parietal junction in patients with schizophrenia. Schizophr Bull. 2016;42:318–26.

16. Jiang J, Zhao K, Li W, Zheng P, Jiang S, Ren Q, et al. Multiomics reveals biological mechanisms linking macroscale structural covariance network dysfunction with neuropsychiatric symptoms across the alzheimer’s disease continuum. Biol Psychiatry. 2025;97:1067–78.

17. Hansen JY, Shafiei G, Markello RD, Smart K, Cox SML, Nørgaard M, et al. Mapping neurotransmitter systems to the structural and functional organization of the human neocortex. Nat Neurosci. 2022;25:1569–81.

18. Foss-Feig JH, Adkinson BD, Ji JL, Yang G, Srihari VH, McPartland JC, et al. Searching for cross-diagnostic convergence: Neural mechanisms governing excitation and inhibition balance in schizophrenia and autism spectrum disorders. Biol Psychiatry. 2017;81:848–61.

19. Lisman J. Excitation, inhibition, local oscillations, or large-scale loops: What causes the symptoms of schizophrenia? Curr Opin Neurobiol. 2012;22:537–44.

20. Wulff K, Gatti S, Wettstein JG, Foster RG. Sleep and circadian rhythm disruption in psychiatric and neurodegenerative disease. Nat Rev Neurosci. 2010;11:589–99.

21. Deco G, Ponce-Alvarez A, Hagmann P, Romani GL, Mantini D, Corbetta M. How local excitation-inhibition ratio impacts the whole brain dynamics. J Neurosci. 2014;34:7886–98.

22. Zhang H, Huang X, Wang C, Liang K. Alteration of gamma-aminobutyric acid in the left dorsolateral prefrontal cortex of individuals with chronic insomnia: A combined transcranial magnetic stimulation-magnetic resonance spectroscopy study. Sleep Med. 2022;92:34–40.

23. Dukart J, Holiga S, Rullmann M, Lanzenberger R, Hawkins PCT, Mehta MA, et al. Juspace: A tool for spatial correlation analyses of magnetic resonance imaging data with nuclear imaging derived neurotransmitter maps. Hum Brain Mapp. 2021;42:555–66.

24. Liu J, Xia M, Wang X, Liao X, He Y. The spatial organization of the chronnectome associates with cortical hierarchy and transcriptional profiles in the human brain. Neuroimage. 2020;222:117296.

25. Chu T, Si X, Xie H, Ma H, Shi Y, Yao W, et al. Regional structural-functional connectivity coupling in major depressive disorder is associated with neurotransmitter and genetic profiles. Biol Psychiatry. 2025;97:290–301.

26. Zarkali A, McColgan P, Ryten M, Reynolds R, Leyland LA, Lees AJ, et al. Differences in network controllability and regional gene expression underlie hallucinations in parkinson’s disease. Brain. 2020;143:3435–48.

27. Fiore A, Preziosa P, Tedone N, Margoni M, Vizzino C, Mistri D, et al. Correspondence among gray matter atrophy and atlas-based neurotransmitter maps is clinically relevant in multiple sclerosis. Mol Psychiatry. 2023;28:1770–82.

28. Morais RF, Sousa JM, Koba C, Andres L, Jesus T, Baldeiras I, et al. Differential involvement of neurotransmitter pathways in ad, bvftd and mci: Whole-brain mri analysis. Neurobiol Dis. 2025;209:106897.

29. Ren J, Yan L, Zhou H, Pan C, Xue C, Wu J, et al. Unraveling neurotransmitter changes in de novo gba-related and idiopathic parkinson’s disease. Neurobiol Dis. 2023;185:106254.

30. Faul F, Erdfelder E, Lang AG, Buchner A. G*power 3: A flexible statistical power analysis program for the social, behavioral, and biomedical sciences. Behav Res Methods. 2007;39:175–91.

31. Zhou Q, Liu Z, Yu C, Wang Q, Zhuang W, Tang Y, et al. Effect of combined treatment with transcranial direct current stimulation and repetitive transcranial magnetic stimulation compared to monotherapy for the treatment of chronic insomnia: A randomised, double-blind, parallel-group, controlled trial. BMC Med. 2024;22:538.

32. Klem GH, Lüders HO, Jasper HH, Elger C. The ten-twenty electrode system of the international federation. The international federation of clinical neurophysiology. Electroencephalogr Clin Neurophysiol Suppl. 1999;52:3–6.

33. Lee HS, Schreiner L, Jo SH, Sieghartsleitner S, Jordan M, Pretl H, et al. Individual finger movement decoding using a novel ultra-high-density electroencephalography-based brain-computer interface system. Front Neurosci. 2022;16:1009878.

34. Manzar MD, Jahrami HA, Bahammam AS. Structural validity of the insomnia severity index: A systematic review and meta-analysis. Sleep Med Rev. 2021;60:101531.

35. Morin CM, Belleville G, Bélanger L, Ivers H. The insomnia severity index: Psychometric indicators to detect insomnia cases and evaluate treatment response. Sleep. 2011;34:601–8.

36. Bastien CH, Vallières A, Morin CM. Validation of the insomnia severity index as an outcome measure for insomnia research. Sleep Med. 2001;2:297–307.

37. Wang J, Wang X, Xia M, Liao X, Evans A, He Y. Gretna: A graph theoretical network analysis toolbox for imaging connectomics. Front Hum Neurosci. 2015;9:386.

38. Nørgaard M, Beliveau V, Ganz M, Svarer C, Pinborg LH, Keller SH, et al. A high-resolution in vivo atlas of the human brain’s benzodiazepine binding site of gaba(a) receptors. Neuroimage. 2021;232:117878.

39. Alakurtti K, Johansson JJ, Joutsa J, Laine M, Bäckman L, Nyberg L, et al. Long-term test-retest reliability of striatal and extrastriatal dopamine d2/3 receptor binding: Study with [(11)c]raclopride and high-resolution pet. J Cereb Blood Flow Metab. 2015;35:1199–205.

40. Dukart J, Holiga Š, Chatham C, Hawkins P, Forsyth A, McMillan R, et al. Cerebral blood flow predicts differential neurotransmitter activity. Sci Rep. 2018;8:4074.

41. Savli M, Bauer A, Mitterhauser M, Ding YS, Hahn A, Kroll T, et al. Normative database of the serotonergic system in healthy subjects using multi-tracer pet. Neuroimage. 2012;63:447–59.

42. Arnatkeviciute A, Fulcher BD, Fornito A. A practical guide to linking brain-wide gene expression and neuroimaging data. Neuroimage. 2019;189:353–67.

43. Li J, Seidlitz J, Suckling J, Fan F, Ji GJ, Meng Y, et al. Cortical structural differences in major depressive disorder correlate with cell type-specific transcriptional signatures. Nat Commun. 2021;12:1647.

44. Gandal MJ, Haney JR, Parikshak NN, Leppa V, Ramaswami G, Hartl C, et al. Shared molecular neuropathology across major psychiatric disorders parallels polygenic overlap. Science. 2018;359:693–97.

45. Chen J, Bardes EE, Aronow BJ, Jegga AG. Toppgene suite for gene list enrichment analysis and candidate gene prioritization. Nucleic Acids Res. 2009;37:W305–11.

46. Xu X, Wells AB, O’Brien DR, Nehorai A, Dougherty JD. Cell type-specific expression analysis to identify putative cellular mechanisms for neurogenetic disorders. J Neurosci. 2014;34:1420–31.

47. Bassett DS, Wymbs NF, Porter MA, Mucha PJ, Carlson JM, Grafton ST. Dynamic reconfiguration of human brain networks during learning. Proc Natl Acad Sci U S A. 2011;108:7641–6.

48. Newman ME, Girvan M. Finding and evaluating community structure in networks. Phys Rev E Stat Nonlin Soft Matter Phys. 2004;69:026113.

49. Rosvall M, Bergstrom CT. Maps of random walks on complex networks reveal community structure. Proc Natl Acad Sci U S A. 2008;105:1118–23.

50. Peng W, Wu Z, Song K, Zhang S, Li Y, Xu M. Regulation of sleep homeostasis mediator adenosine by basal forebrain glutamatergic neurons. Science. 2020;369.

51. Malinow R, Malenka RC. Ampa receptor trafficking and synaptic plasticity. Annu Rev Neurosci. 2002;25:103–26.

52. Traynelis SF, Wollmuth LP, McBain CJ, Menniti FS, Vance KM, Ogden KK, et al. Glutamate receptor ion channels: Structure, regulation, and function. Pharmacol Rev. 2010;62:405–96.

53. Paus T, Keshavan M, Giedd JN. Why do many psychiatric disorders emerge during adolescence? Nat Rev Neurosci. 2008;9:947–57.

54. Shi S, Hayashi Y, Esteban JA, Malinow R. Subunit-specific rules governing ampa receptor trafficking to synapses in hippocampal pyramidal neurons. Cell. 2001;105:331–43.

55. Lau CG, Zukin RS. Nmda receptor trafficking in synaptic plasticity and neuropsychiatric disorders. Nat Rev Neurosci. 2007;8:413–26.

56. Paoletti P, Bellone C, Zhou Q. Nmda receptor subunit diversity: Impact on receptor properties, synaptic plasticity and disease. Nat Rev Neurosci. 2013;14:383–400.

57. Coyle JT, Ruzicka WB, Balu DT. Fifty years of research on schizophrenia: The ascendance of the glutamatergic synapse. Am J Psychiatry. 2020;177:1119–28.

58. Mohn AR, Gainetdinov RR, Caron MG, Koller BH. Mice with reduced nmda receptor expression display behaviors related to schizophrenia. Cell. 1999;98:427–36.

59. Hardingham GE, Bading H. Synaptic versus extrasynaptic nmda receptor signalling: Implications for neurodegenerative disorders. Nat Rev Neurosci. 2010;11:682–96.

60. Zhou Y, Won J, Karlsson MG, Zhou M, Rogerson T, Balaji J, et al. Creb regulates excitability and the allocation of memory to subsets of neurons in the amygdala. Nat Neurosci. 2009;12:1438–43.

